# Association of aortic clamping time with systemic immune inflammation and systemic inflammatory response indexes in isolated coronary bypass surgery

**DOI:** 10.1101/2024.04.22.24306198

**Authors:** Duygu Durmaz, Sedat Gündöner, Hayrettin Tekümit

## Abstract

**Objective:** Aortic clamping time during cardiopulmonary bypass (CPB) has been associated with inflammatory processes such as systemic inflammatory response syndrome. In this study, we evaluated the association of CPB and aortic cross-clamping (ACC) times with systemic inflammatory response index (SIRI) and systemic immune inflammation index (SIII) in isolated coronary artery bypass surgery (CABG).

**Methods:** 96 patients who underwent isolated CABG at a single center between 2021 and 2023 were retrospectively analyzed. Patients were divided into below median aortic clamp time (group I; n=56) and above median aortic clamp time (group II; n=42) according to median aortic clamp time (66.2 minutes). Demographic data, preoperative and postoperative laboratory parameters were recorded. SIRI and SIII values were calculated.

**Results:** Baseline demographic data were similar between the groups. The duration of CPB and aortic clamping was significantly longer in group II (p<0.001). SIII and SIRI values were significantly increased in both groups in the postoperative period. However, there was no correlation between increased CPB and ACC durations and SIII and SIRI. However, no significant difference was observed in postoperative SIII and SIRI values between the groups. A weak correlation was found between SIII index and postoperative albumin levels.

**Conclusions:** There is no significant relationship between aortic clamping time and inflammatory indices in patients undergoing isolated CABG surgery. Increasing the duration of surgery does not affect the change in SIRI and SIII values.

## 1. Introduction

During cardiopulmonary bypass (CPB), the patient receives constant blood flow through a mechanical pump (1). In CPB, a cross-clamp is placed over the aorta to provide an immobilized and bloodless operating field (2). Systemic inflammatory response syndrome (SIRS) is an inflammatory process that can be triggered by cardiac surgery and CPB. SIRS is associated with the release of several proinflammatory mediators after open heart surgery procedures that may affect postoperative outcomes (3).

The marked increase in SIRS after coronary artery bypass grafting (CABG) may be related to the duration of perioperative CPB and aortic cross clamping (ACC). Many studies have shown that the duration of CPB and ACC during cardiac surgery are predictors of mortality and morbidity. However, there is no consensus on the safe time limit. In this regard, researchers have reported different results for the safe use of CPB and ACC (4,5).

The concepts of systemic inflammatory response index (SIRI) and systemic immune inflammation index (SIII) have been defined as new inflammatory markers (6,7). These indices include the main components of inflammatory markers such as neutrophils, monocytes, lymphocytes and platelets and have been presented as possible mortality prognostic factors in different cardiovascular diseases (8,9).

In this study, we tried to determine the relationship between aortic clamping time and inflammatory indexes in isolated coronary artery bypass surgery.

## 2. Methods

### 2.1 Study Design and Patients

The population of this retrospective study consisted of 96 patients undergoing first-time isolated CABG at a single center between June 2021 and September 2023. Patients over 18 years of age who underwent elective isolated on-pump coronary artery bypass surgery were included. Reoperations, patients with additional surgical procedures, patients with known cerebrovascular disease, bleeding pathology and patients with incomplete medical data were excluded. The study groups were divided into those with a median aortic clamping time of the total population below (Group I) and above (Group II) 66.2 minutes. The study was approved by XXX University Non-Interventional Research Ethics Committee with the ethics committee decision dated 16.01.2024 and numbered 2024-1.

### 2.2 Data collection and calculation of inflammatory indexes

Demographic data, preoperative and postoperative hematologic and biochemical analyses, and clinical data including postoperative processes were obtained by examining the computerized patient record system. SIRI and SIII indexes related with systemic inflammation were calculated as follows (10). SIII; peripheral platelet count x neutrophil count / lymphocyte count. SIRI; neutrophil count x monocyte count / lymphocyte count

### 2.3 Operative technique

All patients underwent standard median sternotomy under general anesthesia. After appropriate anticoagulation with 400 IU/kg heparin, cardiopulmonary bypass (CPB) was initiated by cannulation of the ascending aorta and right atrium. All patients were monitored with moderate hypothermia (28 - 32 °C) and myocardial protection was achieved with antegrade-retrograde combined blood cardioplegia. Mean arterial pressure was maintained between 60 and 80 mmHg during CPB. After the procedure was completed, CPB was terminated after the body temperature of the patients was raised to 37 °C. Decanulation was performed after neutralization with protamine.

### 2.4 Statistical analysis

Statistical evaluation was performed with IBM SPSS 25.0 (SPSS Inc., Chicago, IL, USA) package program. While evaluating the study data, quantitative variables were represented by mean, standard deviation, median, min and max values, and qualitative variables were represented by descriptive statistical methods such as frequency and percentage. Shapiro Wilks test and Box Plot graphs were used to evaluate the conformity of the data to normal distribution. Student’s t-test was used for quantitative two-group evaluations showing normal distribution, and Mann Whitney-U test was used for those not showing normal distribution. Between two follow-ups, Paired Samples test was used for those with normal distribution and Wilcoxon test was used for those without normal distribution. Chi-Square test and Fisher Exact Test were used to compare qualitative data. Pearson correlation analysis was used to evaluate the relationships between variables. The results were evaluated at 95% confidence interval and significance was evaluated at p<0.05 level.

## 3. Results

A total of 96 patients who underwent isolated CABG were included in the study. The mean ages of the patients included in the study were 63.6±8.8 years for group I and 66±8.9 years for group II (p>0.05). 64.8% of patients in group I and 73.8% of patients in group II were male. The study groups were divided into those with a median aortic clamping time of the total population below (Group I) and above (Group II) 66.2 minutes. The mean aortic clamping times were 51.7±10.7 minutes for group I and 89.9±13.0 minutes for group II (p<0.05) (Table 1).

**Table 1:**
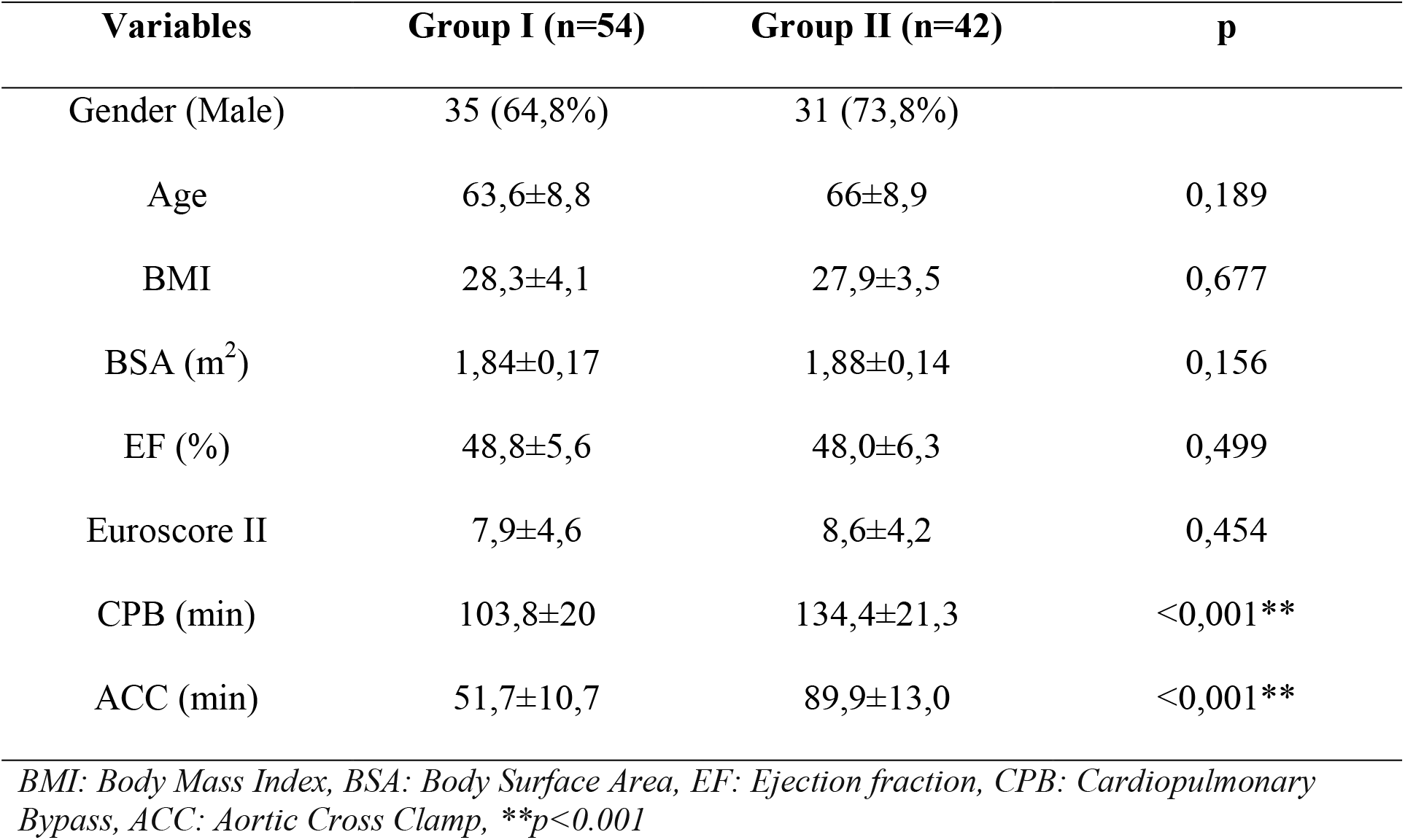
Demographic and operative data of patients.

The changes in the parameters within and between the groups at preoperative and postoperative 24th hour periods are shown in Table 2. Significant differences were found between group I and group II in SIII, SIRI, CRP and albumin values at preoperative and postoperative 24 hours. In the comparison between the groups, preoperative and postoperative SIII, SIRI and albumin values were similar. Although there was a statistical difference in preoperative CRP levels between the groups, no statistical difference was found between the CRP values in the postoperative 24th hour measurements (Table 2). The graphical changes of SIII and SIRI parameters of group I and group II are shown in figures I and II.

**Figure 1:**
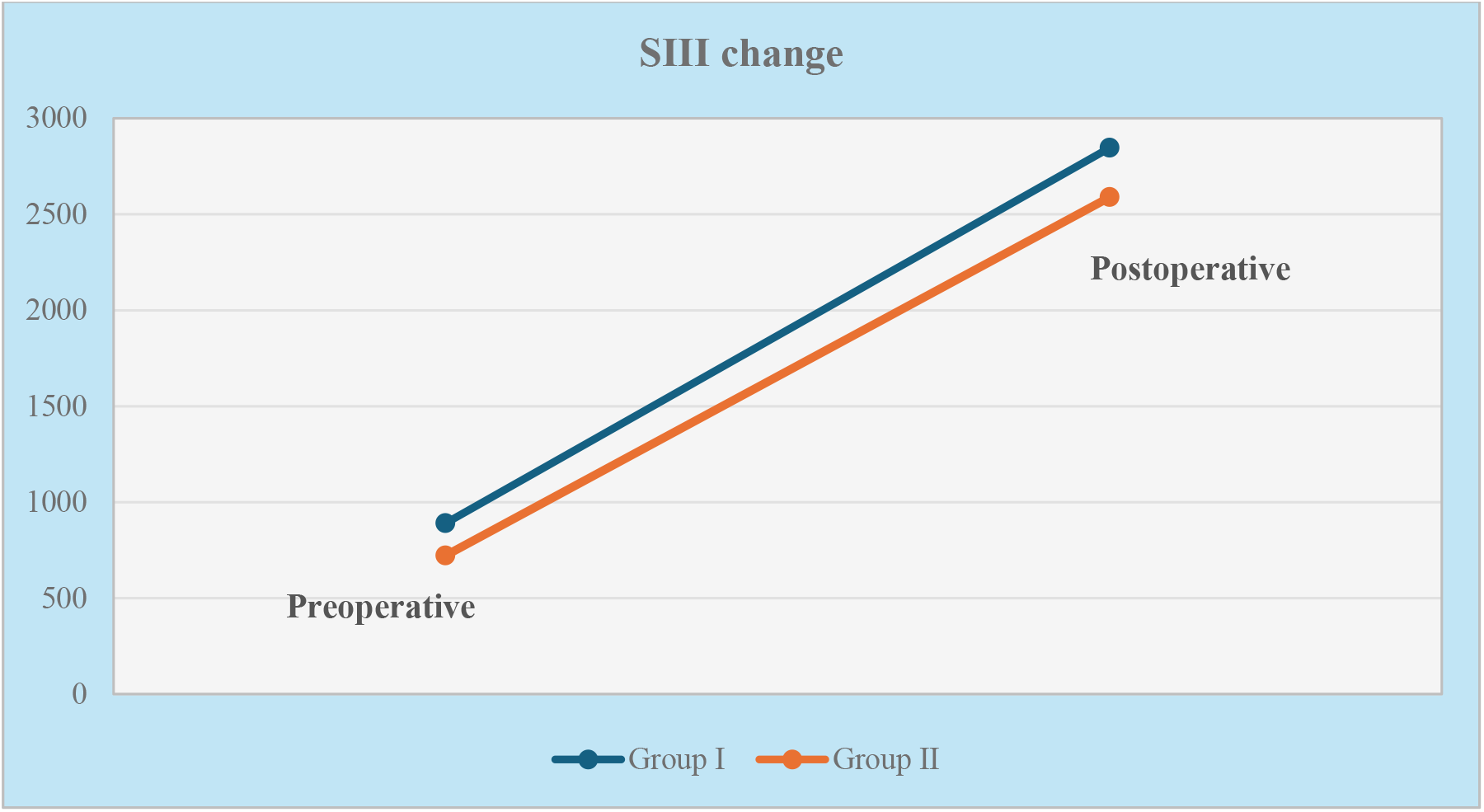
SIII change according to groups.

**Figure 2:**
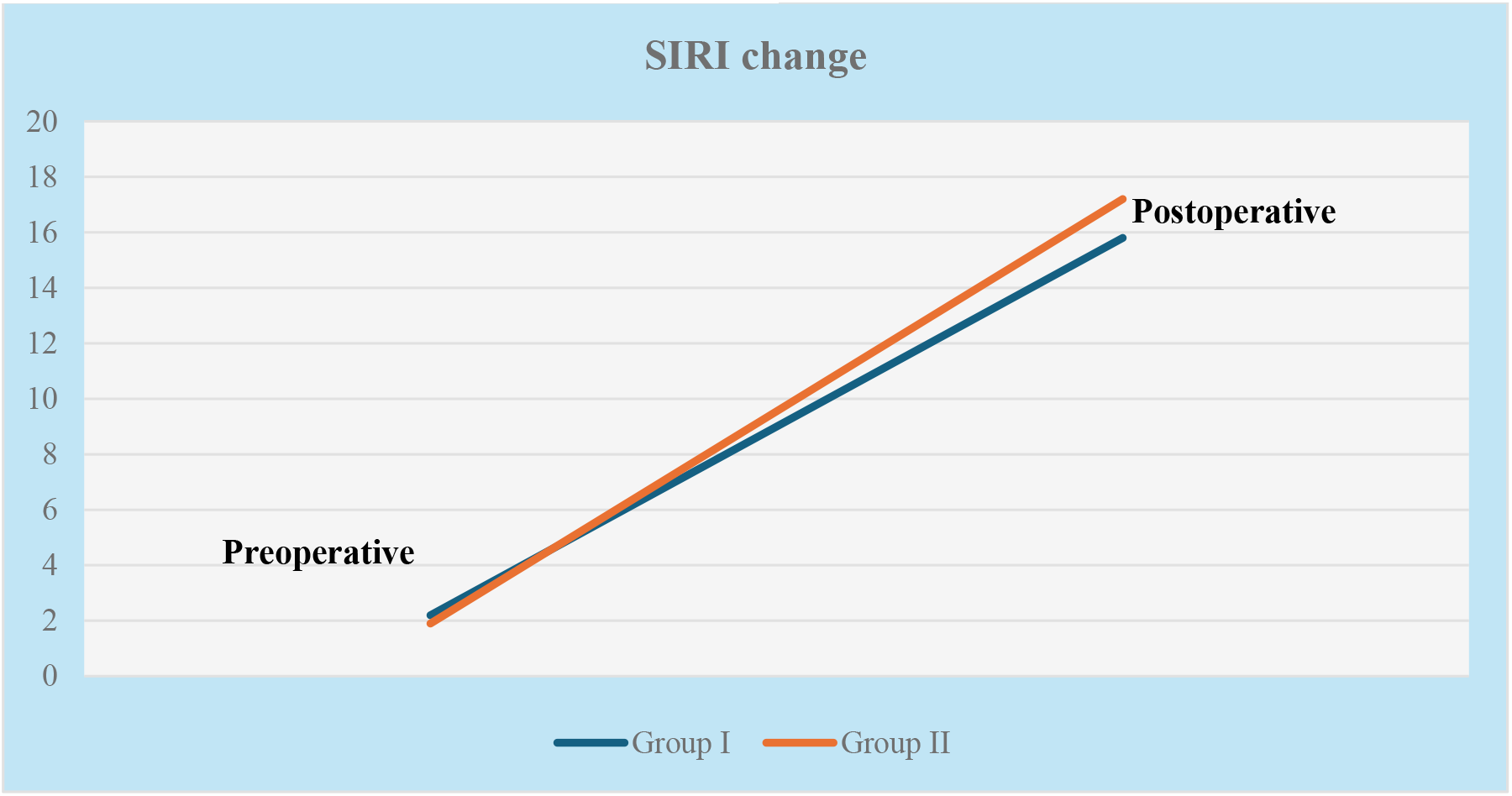
SIRI change according to groups.

**Table 2:** Preoperative and Postoperative Laboratory Parameters.

|  |  | Group |  | *p* |
| --- | --- | --- | --- | --- |
|  |  | Group I (n=54) | Group II (n=42) |  |
| SIII |  |  |  |  |
| Preoperative | Mean±SD | 890,8±680,2 | 721,7±417,5 | <sup>d</sup>0,404 |
| Postoperative | Mean±SD | 2847,3±1626,6 | 2590,3±1354,1 | <sup>d</sup>0,07 |
|  | <sup>e</sup>*p* | 0,001** | 0,001** |  |
| SIRI |  |  |  |  |
| Preoperative | Mean±SD | 2,2±1,5 | 1,9±1,3 | <sup>d</sup>0,520 |
| Postoperative | Mean±SD | 15,8±9,4 | 17,2±9,8 | <sup>d</sup>0,398 |
|  | <sup>e</sup>*p* | 0,001** | 0,001** |  |
| Albümin |  |  |  |  |
| Preoperative | Mean±SD | 4,1±0,4 | 4,2±0,4 | <sup>b</sup>0,224 |
| Postoperative | Mean±SD | 3,5±0,3 | 3,4±0,4 | <sup>d</sup>0,311 |
|  | <sup>e</sup>*p* | 0,001** | 0,001** |  |
| CRP |  |  |  |  |
| Preoperative | Mean±SD | 2,2±2,3 | 3,0±1,2 | <sup>d</sup>0,001** |
| Postoperative | Mean±SD | 9,2±2,8 | 8,0±3,9 | <sup>b</sup>0,104 |
|  | <sup>e</sup>*p* | 0,001** | 0,001** |  |

*SII*; systemic immun inflammation index, *SIRI*; systemic immun response index, *CRP*; C-reactive protein, <sup>b</sup>Student-t Test, <sup>d</sup>Mann-Whitney-U Test, <sup>e</sup>Paired Samples t-Test, \*\**p*<0,001

When the correlation of SIRI and SIII values with CPB and ACC duration, postoperative albumin and CRP values were analyzed, it was found that only postoperative SIII value had a weak correlation with postoperative albumin level (r=0.240; p=0.019) (Table 3). No statistically significant correlation was found for other postoperative variables.

**Table 3:**
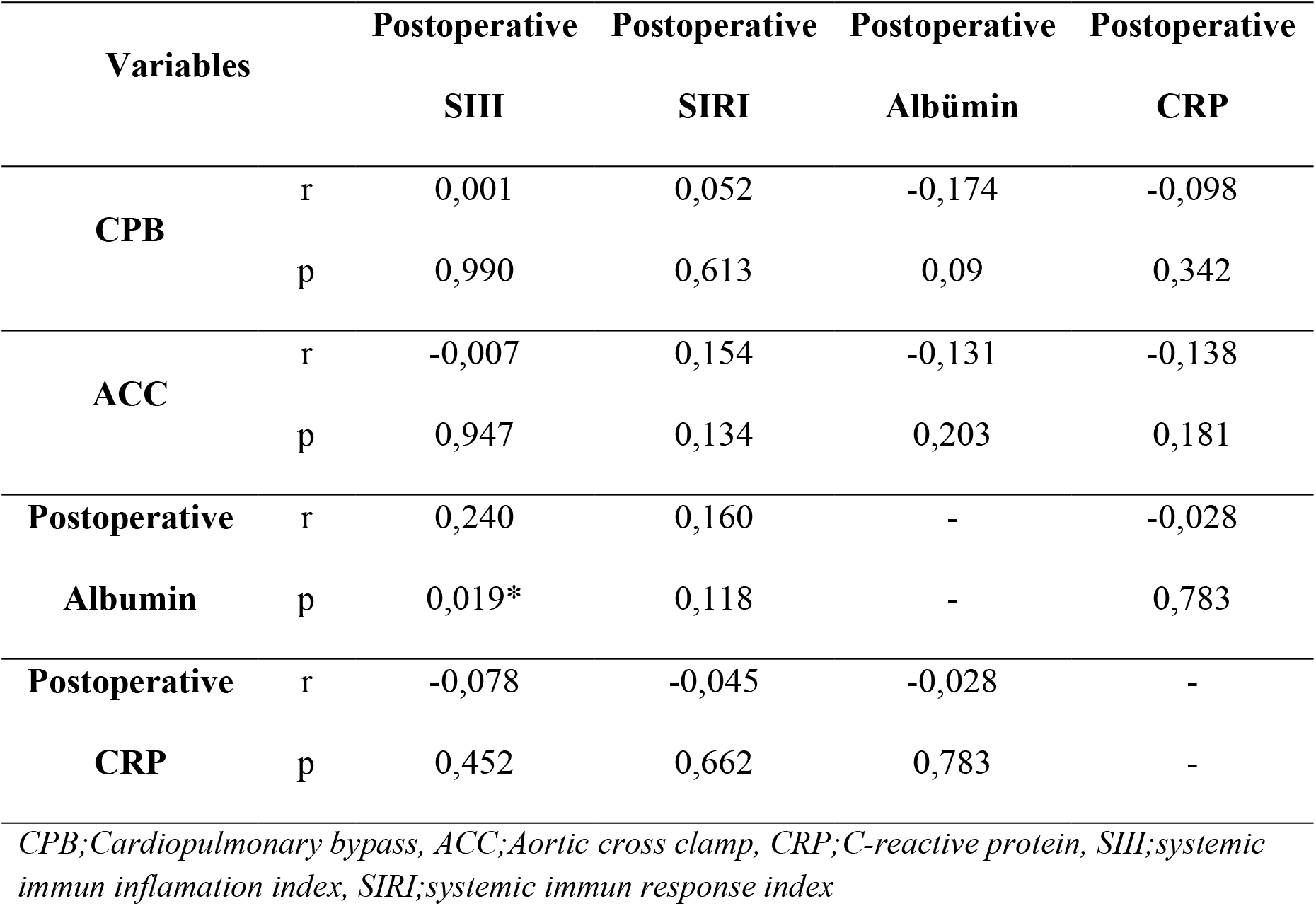
Correlation between variables.

## 4. Discussion

To our knowledge, this is the first study to present the relationship and variation of different inflammatory indices with aortic cross-clamping time in on-pump coronary artery bypass surgery. In this retrospective study, we evaluated the relationship between aortic clamp duration and inflammatory indices in 96 patients undergoing isolated CABG surgery. Different studies have shown that some complications develop and the risk of death increases in patients exposed to prolonged aortic cross-clamping (11,12,13).

The systemic immune inflammatory index (SIII) is an inflammatory index that includes three differently shaped elements involved in the immune response, including neutrophils, lymphocytes and platelets. This index has been reported to be a prognostic marker used to determine long-term prognosis in coronary revascularization (14). Researchers have also described this index as an important marker reflecting the inflammatory status in various clinical pictures including appendicitis, coronary artery disease and neoplasms (15,16,17).

Systemic inflammatory response describes the pathophysiologic response of the immune system to events such as infection, trauma, burns and various other injuries (18). Many factors associated with the contact of blood with extracorporeal artificial circuits in cardiac surgery contribute to SIRS formation (19). The systemic inflammatory response index (SIRI) is defined as an inflammatory index describing immunologic defenses involving neutrophils, monocytes and lymphocytes (20).

Although CPB is considered a safe method to facilitate cardiac surgery, prolonged CPB is associated with complications (21). Exposure of blood to abnormal shear stress and contact with the artificial surface of the bypass circuit leads to pro-inflammatory activation of the coagulation and complement systems, endothelial cell death and platelet activation (22,23,24). In addition, prolonged aortic cross-clamping time is known to be associated with decreased early survival after cardiac surgery (25). It is not known how the inflammatory indexes SIII and SIRI, which have emerged as a prognostic factor for cardiovascular diseases, change depending on the duration of CPB and aortic cross-clamping. In our study, we tried to determine the effect of different aortic cross-clamping times on these indexes in patients undergoing isolated on-pump CABG.

We divided the included patients into two groups according to the median aortic cross-clamp time (66.2 min) of patients undergoing isolated CABG. The aortic cross-clamp time was 51.7±10.7 and 89.9±13.0 min in group I and group II, respectively. Baseline demographic characteristics of the patients were similar. Mean CPB duration was 103.8±20 and 134.4±21.3 min in group I and group II, respectively. Preoperative SIII and SIRI values were similar in both groups. In the postoperative period, there was an increase in these indices within the groups. We think that this is related to the increase in the inflammatory response to CPB. However, postoperative SIII and SIRI indices were similar between the groups. This suggests that the relatively long aortic clamping time did not affect the inflammatory indices. Robic M et al. reported that a longer duration of CPB was associated with increased blood neutrophil count (26). In our study, it was determined that the SIII value including the neutrophil parameter was not related with duration.

C-Reactive Protein (CRP) increases significantly in response to inflammation. CRP levels increase significantly after cardiac surgery with CPB (27). In our study, although postoperative CRP values increased statistically significantly in both groups compared to preoperative values, postoperative measurements were similar between the groups. In cardiac surgery with CPB, serum albumin levels decrease with the effect of hemodilution (28). At the same time, high inflammation parameters have been independently associated with hypoalbuminemia (29). In our study, a significant decrease in postoperative serum albumin levels was observed in both groups and postoperative serum albumin levels were similar between the groups.

There was no correlation between postoperative SIII and SIRI indices and CPB duration, aortic clamping duration and CRP levels. However, a weak statistically significant correlation was found between postoperative SIII index and postoperative albumin levels (r=0.252; p=0.018).

Our study is the first to evaluate the relationship between inflammatory indices and operative times in patients undergoing isolated CABG surgery. However, there are some limitations. First of all, our study has a non-randomized design because it is retrospective. This creates the possibility of limiting the population included. In addition, the fact that it is a single-center study also limits the sample size. We believe that the relationship between CPB and inflammatory indices should be investigated in detail with a larger sample group.

## Data Availability

The availability of data for all citations in this article is based on information taken from relevant sources and believed to be reliable.

## Conflict of interest

None

## Funding

None

## Acknowledgements

None

## Notes

### Competing Interest Statement

The authors have declared no competing interest.

### Clinical Trial

This is a retrospectively designed study

### Funding Statement

there is no funding support in this text

### Author Declarations

BANDIRMA 17 EYLÜL UNIVERSITY RECTORATE HEALTH SCIENCES NON-INTERVENTIONAL RESEARCH ETHICS COMMITTEE

## References

1) Sarkar M, Prabhu V. Basics of cardiopulmonary bypass. Indian J Anaesth. 2017 Sep;61(9):760–767. doi: 10.4103/ija.IJA_379_17. PMID: 28970635; PMCID: PMC5613602.

2) Machin D, Allsager C. Principles of cardiopulmonary bypass. Contin Educ Anaesth Crit Care Pain. 2006;6(5):176–181. doi: 10.1093/bjaceaccp/mkl043.

3) Gümüs F, Erkalp K, Kayalar N, Alagöl A. Yasli hasta popülasyonunda kalp cerrahisi ve anestezi yaklasimi. Türk Gögüs Kalp Damar Dergisi. 2013; 21 (1):250–255.

4) Verheijen LP, van Zaane B, van Aarnhem EE, Peelen LM, van Klei WA. The association between aortic cross clamp time and postoperative morbidity and mortality in mitral valve repair: a retrospective cohort study. J Cardiovasc Surg (Torino). 2018 Jun;59(3):453–461. doi: 10.23736/S0021-9509.18.10123-6. Epub 2018 Feb 8. PMID: 29430884.

5) Mehmood A, Nadeem RN, Kabbani MS, Khan AH, Hijazi O, Ismail SR, Shath G, Eng WW, Jawed S. Impact of Cardiopulmonary Bypass and Aorta Cross Clamp Time on the Length of Mechanical Ventilation after Cardiac Surgery among Children: A Saudi Arabian Experience. Cureus. 2019 Aug 7;11(8):e5333. doi: 10.7759/cureus.5333. PMID: 31598440; PMCID: PMC6778048.

6) Parmana IMA, Boom CE, Poernomo H, Gani C, Nugroho B, Cintyandy R, Sanjaya L, Hadinata Y, Parna DR, Hanafy DA. Systemic Immune-Inflammation Index Predicts Prolonged Mechanical Ventilation and Intensive Care Unit Stay After off-Pump Coronary Artery Bypass Graft Surgery: A Single-Center Retrospective Study. Vasc Health Risk Manag. 2023;19:353–361 10.2147/VHRM.S409678

7) Urbanowicz T, Michalak M, Olasinska-Wisniewska A, Rodzki M, Witkowska A, Gasecka A, Buczkowski P, Perek B, Jemielity M. Neutrophil Counts, Neutrophil-to-Lymphocyte Ratio, and Systemic Inflammatory Response Index (SIRI) Predict Mortality after Off-Pump Coronary Artery Bypass Surgery. Cells. 2022 Mar 26;11(7):1124. doi: 10.3390/cells11071124. PMID: 35406687; PMCID: PMC8997598.

8) Peng Y, Huang W, Shi Z, Chen Y, Ma J. Positive association between systemic immune-inflammatory index and mortality of cardiogenic shock. Clin Chim Acta. 2020 Dec;511:97–103. doi: 10.1016/j.cca.2020.09.022. Epub 2020 Oct 9. PMID: 33045194.

9) Luo H, He L, Zhang G, Yu J, Chen Y, Yin H, Goyal H, Zhang GM, Xiao Y, Gu C, Yin M, Jiang X, Song X, Zhang L. Normal Reference Intervals of Neutrophil-To-Lymphocyte Ratio, Platelet-To-Lymphocyte Ratio, Lymphocyte-To-Monocyte Ratio, and Systemic Immune Inflammation Index in Healthy Adults: a Large Multi-Center Study from Western China. Clin Lab. 2019 Mar 1;65(3). doi: 10.7754/Clin.Lab.2018.180715. PMID: 30868857.

10) Lin KB, Fan FH, Cai MQ, Yu Y, Fu CL, Ding LY, Sun YD, Sun JW, Shi YW, Dong ZF, Yuan MJ, Li S, Wang YP, Chen KK, Zhu JN, Guo XW, Zhang X, Zhao YW, Li JB, Huang D. Systemic immune inflammation index and system inflammation response index are potential biomarkers of atrial fibrillation among the patients presenting with ischemic stroke. Eur J Med Res. 2022 Jul 2;27(1):106. doi: 10.1186/s40001-022-00733-9. PMID: 35780134; PMCID: PMC9250264.

11) Torsten Doenst, Michael A. Borger, Richard D. Weisel, Terrence M. Yau, Manjula Maganti, Vivek Rao, Relation between aortic cross-clamp time and mortality — not as straightforward as expected, European Journal of Cardio-Thoracic Surgery, Volume 33, Issue 4, April 2008, Pages 660–665, 10.1016/j.ejcts.2008.01.001

12) Argyris Michalopoulos, George Tzelepis, Urania Dafni, Stefanos Geroulanos, Determinants of Hospital Mortality After Coronary Artery Bypass Grafting, Chest, Volume 115, Issue 6, 1999, Pages 1598–1603, ISSN 0012-3692, 10.1378/chest.115.6.1598.

13) Jeffrey P. Schwartz, Mamdouh Bakhos, Amit Patel, Sally Botkin, Siyamek Neragi-Miandoab, Repair of aortic arch and the impact of cross-clamping time, New York Heart Association stage, circulatory arrest time, and age on operative outcome, Interactive CardioVascular and Thoracic Surgery, Volume 7, Issue 3, June 2008, Pages 425–429, 10.1510/icvts.2007.164871

14) Urbanowicz TK, Michalak M, Gasecka A, Olasinska-Wisniewska A, Perek B, Rodzki M, Bociański M, Jemielity M. A Risk Score for Predicting Long-Term Mortality Following Off-Pump Coronary Artery Bypass Grafting. J Clin Med. 2021 Jul 7;10(14):3032. doi: 10.3390/jcm10143032. PMID: 34300198; PMCID: PMC8305554.

15) Hajibandeh S, Hajibandeh S, Hobbs N, Mansour M. Neutrophil-to-lymphocyte ratio predicts acute appendicitis and distinguishes between complicated and uncomplicated appendicitis: A systematic review and meta-analysis. Am J Surg. 2020 Jan;219(1):154–163. doi: 10.1016/j.amjsurg.2019.04.018. Epub 2019 Apr 27. PMID: 31056211.

16) Aydin C, Engin M. The Value of Inflammation Indexes in Predicting Patency of Saphenous Vein Grafts in Patients With Coronary Artery Bypass Graft Surgery. Cureus. 2021 Jul 26;13(7):e16646. doi: 10.7759/cureus.16646. PMID: 34462681; PMCID: PMC8387011.

17) Li W, Ma G, Deng Y, Chen W, Liu Z, Chen F, Wu Q. Systemic Immune-Inflammation Index Is a Prognostic Factor for Breast Cancer Patients After Curative Resection. Front Oncol. 2021 Dec 1;11:570208. doi: 10.3389/fonc.2021.570208. PMID: 34926234; PMCID: PMC8671143.

18) Balk RA. Systemic inflammatory response syndrome (SIRS): where did it come from and is it still relevant today? Virulence. 2014 Jan 1;5(1):20–6. doi: 10.4161/viru.27135. Epub 2013 Nov 13. PMID: 24280933; PMCID: PMC3916374.

19) Day JR, Taylor KM. The systemic inflammatory response syndrome and cardiopulmonary bypass. Int J Surg. 2005;3(2):129–40. doi: 10.1016/j.ijsu.2005.04.002. Epub 2005 Aug 1. PMID: 17462274.

20) Urbanowicz T, Michalak M, Olasinska-Wisniewska A, Rodzki M, Witkowska A, Gąsecka A, Buczkowski P, Perek B, Jemielity M. Neutrophil Counts, Neutrophil-to-Lymphocyte Ratio, and Systemic Inflammatory Response Index (SIRI) Predict Mortality after Off-Pump Coronary Artery Bypass Surgery. Cells. 2022 Mar 26;11(7):1124. doi: 10.3390/cells11071124. PMID: 35406687; PMCID: PMC8997598.

21) Salis S, Mazzanti VV, Merli G, Salvi L, Tedesco CC, Veglia F and Sisillo E. Cardiopulmonary Bypass Duration Is an Independent Predictor of Morbidity and Mortality After Cardiac Surgery. Journal of Cardiothoracic and Vascular Anesthesia. 2008;22:814–822.

22) Schmid FX, Vudattu N, Floerchinger B, Hilker M, Eissner G, Hoenicka M, Holler E and Birnbaum DE. Endothelial apoptosis and circulating endothelial cells after bypass grafting with and without cardiopulmonary bypass. Eur J Cardiothorac Surg. 2006;29:496–500.

23) Verrier ED and Morgan EN. Endothelial response to cardiopulmonary bypass surgery. Ann Thorac Surg. 1998;66:S17–9; discussion S25–8.

24) Weerasinghe A and Taylor KM. The platelet in cardiopulmonary bypass. Ann Thorac Surg. 1998;66:2145–52.

25) Iino K, Miyata H, Motomura N, Watanabe G, Tomita S, Takemura H, Takamoto S. Prolonged Cross-Clamping During Aortic Valve Replacement Is an Independent Predictor of Postoperative Morbidity and Mortality: Analysis of the Japan Cardiovascular Surgery Database. Ann Thorac Surg. 2017 Feb;103(2):602–609. doi: 10.1016/j.athoracsur.2016.06.060. Epub 2016 Sep 10. PMID: 27624296.

26) Robich M, Ryzhov S, Kacer D, Palmeri M, Peterson SM, Quinn RD, Carter D, Sheppard F, Hayes T, Sawyer DB, Rappold J, Prudovsky I, Kramer RS. Prolonged Cardiopulmonary Bypass is Associated With Endothelial Glycocalyx Degradation. J Surg Res. 2020 Jul;251:287–295. doi: 10.1016/j.jss.2020.02.011. Epub 2020 Apr 30. PMID: 32199337; PMCID: PMC7247933.

27) Aouifi A, Piriou V, Blanc P, Bouvier H, Bastien O, Chiari P, Rousson R, Evans R, Lehot JJ. Effect of cardiopulmonary bypass on serum procalcitonin and C-reactive protein concentrations. Br J Anaesth. 1999 Oct;83(4):602–7. doi: 10.1093/bja/83.4.602. PMID: 10673877.

28) Berbel-Franco D, Lopez-Delgado JC, Putzu A, Esteve F, Torrado H, Farrero E, Rodríguez-Castro D, Carrio ML, Landoni G. The influence of postoperative albumin levels on the outcome of cardiac surgery. J Cardiothorac Surg. 2020 May 11;15(1):78. doi: 10.1186/s13019-020-01133-y. PMID: 32393356; PMCID: PMC7216430.

29) Eckart A, Struja T, Kutz A, Baumgartner A, Baumgartner T, Zurfluh S, Neeser O, Huber A, Stanga Z, Mueller B, Schuetz P. Relationship of Nutritional Status, Inflammation, and Serum Albumin Levels During Acute Illness: A Prospective Study. Am J Med. 2020 Jun;133(6):713-722.e7. doi: 10.1016/j.amjmed.2019.10.031. Epub 2019 Nov 18. PMID: 31751531.

